# The impact of the COVID-19 pandemic and related control measures on cancer diagnosis in Catalonia: A time-series analysis of primary care electronic health records covering about 5 million people

**DOI:** 10.1101/2020.11.26.20239202

**Authors:** Ermengol Coma, Carolina Guiriguet, Núria Mora, Mercè Marzo-Castillejo, Mència Benítez, Leonardo Méndez-Boo, Francesc Fina, Mireia Fàbregas, Albert Mercadé, Manuel Medina

## Abstract

**Objectives:** Cancer care has been disrupted by the response of health systems to the COVID-19 pandemic, especially during lockdowns. The aim of our study is to analyse the impact of the pandemic on the incidence of cancer diagnosed in primary care.

**Design:** Time-series study of malignant neoplasm and diagnostic procedures, using data from the primary care electronic health records from January 2014 to September 2020.

**Setting:** Primary care, Catalonia, Spain

**Participants:** People older than 14 years and assigned in one of the primary care practices of the Catalan Institute of Health with a new diagnosis of malignant neoplasm.

**Main outcome measures:** We obtained the monthly expected incidence of malignant neoplasms using a temporary regression, where the response variable was the incidence of cancer from 2014 to 2018 and the adjustment variables were the trend and seasonality of the time series. Excess or lack of malignant neoplasms were defined as the number of observed minus expected cases, globally and stratified by sex, age, type of cancer, and socioeconomic status.

**Results:** Between March and September 2020 we observed 8,766 (95% CI: 4,135 to 13,397) less malignant neoplasm diagnoses, representing a reduction of 34% (95% CI: 19.5% to 44.1%) compared to the expected. This underdiagnosis was greater in individuals aged more than 64 years, men, and in some types of cancers (skin, colorectal, prostate). Although the reduction was predominantly focused during the lockdown, expected figures have not yet been reached (40.5% reduction during the lockdown and 24.3% reduction after that).

**Conclusions:** Reduction on cancer incidences has been observed during and after the lockdown. Urgent policy interventions are necessary to mitigate the indirect effects of COVID-19 pandemic and related control measures on other diseases and some strategies must be designed in order to reduce the underdiagnosis of cancer.

**What is already know in this topic:**

- The COVID-19 pandemic and related control measures have significantly affected medical care worldwide, with effects on cancer diagnosis.
- Non-COVID elective services (healthcare consultations, services, cancer screening programmes) were suspended and have been associated with a reduction in incidence of cancer.
- Skin non-melanoma cancers has been more affected than other type of cancers

**What this study adds?:**

- Provides data from a primary care perspective in a population about 5 million people.
- Underdiagnosis of cancer occurred during the lockdown. However, this reduction extended beyond the end of the lockdown, especially for people older than 64 years, men, and some types of cancer.
- Least deprived areas had greater reductions in cancer diagnoses during the lockdown, but after the lockdown the most deprived areas were those with more underdiagnosis.

## Background

The coronavirus disease 2019 (COVID-19) caused by SARS-CoV-2 virus started as an outbreak in Wuhan (China) [1] and quickly evolved into a global pandemic [2]. The first cases in Europe were identified in France on 24 January 2020 [3] and the first official case in Catalonia (Spain) was reported a month later on 25 February. In a few months, COVID-19 has become one of the greatest public health challenges in recent times. By 5 November 2020, SARS-CoV-2 has infected over 48 million people worldwide and caused more than 1,233,000 deaths [4]. In Catalonia, as of the same date, official figures stated there have been 290,244 confirmed COVID-19 cases and 14,482 deaths [5].

In the absence of a vaccine, a social distancing strategy has been the primary intervention to reduce the spread of the epidemic and COVID-19 related deaths [6]. In many countries, this strategy includes a national lockdown [6]. In Spain, lockdown was adopted on 14 March [7]. During and after the lockdown, governments advocated that patients communicated with their general practitioners by phone, encouraged people to stay at home and work from home, cancelled non-essential health care services, restricted some diagnostic and therapeutic procedures to urgent cases only and scaled down most routine preventive activities [8]. Despite the need for these measures to control the epidemic and reduce COVID-19 mortality [9], many studies point out collateral health damages. In fact, a reduction of preventive care measures, like screening [10] or childhood vaccination [11], diagnosis [12] and control of chronic diseases in primary care [13] were reported. In addition, some studies highlighted an increase of mental illness [14], as well as all-cause mortality [15].

Cancer care has also been disrupted by the response of health systems to the pandemic [16]. Some studies have highlighted the problem of access to and utilisation of cancer services during and after the first wave of SARS-CoV-2 [17-19]. Some examples are the limitation of access to screening programmes, to medical care cancer related visits (both in primary care and in hospitals), to cancer diagnostic tests, the reduction in scheduled cancer surgeries due to limited access to post-surgical critical care units and occupation of hospital beds by patients with COVID-19, a reduction of treatments that could pose a high risk to the patient in the circumstances of the pandemic and modifications of therapeutic guidelines, among others [18, 19]. In some countries, a reduction in cancer diagnostics and cancer encounters was observed [20 - 22]. This led to a reduction in new cancer diagnoses and a delay of cancer treatment [23, 24]. There is evidence that progression of cancers during these delays (even short delays of 3 months for more aggressive cancers) will impact on patient long-term survival [22, 25]. In the UK, substantial increases in the number of avoidable cancer deaths are to be expected as a result of diagnosis delays due to the COVID-19 pandemic [26]. As some projections have estimated that COVID-19 related disruption in health care may last for more than 18 months [22], it is crucial to promptly quantify the effects on cancer diagnosis in any health system in order to mitigate their negative effects.

Considering that the future of the COVID-19 pandemic is currently unknown, we need to ensure that patients continue to receive proper cancer care, in order to minimise the clinical impact of a diagnostic delay due to COVID-19-related dysfunctions. The aim of this study is to analyse and interpret the impact of the COVID-19 pandemic on the incidence of malignant neoplasms through the analysis of primary care electronic health records (EHR) data in Catalonia.

## Methods

### Design and data source

We performed a time-series study of malignant neoplasms case reported diagnoses. Data were extracted from primary care EHR of the *Institut Català de la Salut* (Catalan Institute of Health, ICS, for its Catalan initials). ICS is the main primary care provider in Catalonia. It manages around 75% of all primary care practices (PCP) in the Catalan public health system and covers about 5.8 million people. Its population is highly representative of the population of Catalonia in terms of geographic area, age distribution and gender [27]. In 2005, ICS implemented an universal EHR system for use in primary care known as ECAP (*Estació Clínica d’Atenció Primària*), which is a software system that serves as a repository for structured data on diagnoses (coded according to the International Classification of Diseases 10th revision ICD-10), clinical variables, prescription data, laboratory test results and diagnostic requests. Data from ICS’ primary care EHR, including cancer diagnoses, have been previously validated in several studies [28].

### Participants and study period

We included all patients older than 14 years old with a malignant neoplasm diagnosis registered in the EHR (see ICD-10 codes in supplementary table 1). The study period included all months between January 2014 and September 2020. We divided this period into three for the time-series analysis: training (2014-2018), validation (2019) and analysis (2020). In addition, we separated the year 2020 available months into two periods for the calculation of the reductions of cancer incidence: “lockdown period” from March to June coinciding with the state of alarm in Spain (including the lockdown and different phases of de-escalation) and “post-lockdown period” from July to September, after the state of alarm.

**Table 1.** Baseline characteristics of study population by set: training set (2014-2018), validation set (2019) and analysis set (2020)

| Variable | 2014-2018 | 2019 | 2020* |
| --- | --- | --- | --- |
| Population older than 14 years | 4,781,835 | 4,846,503 | 4,927,406 |
| % of women | 51.21 | 51.20 | 51.15 |
| % of population older than 64 years | 22.29 | 22.36 | 22.39 |
| Socioeconomic status: % of people in first quartile (least deprived) | 21.69 | 21.71 | 21.70 |
| Socioeconomic status: % of people in 2nd quartile | 15.23 | 15.23 | 15.25 |
| Socioeconomic status: % of people in 3rd quartile | 20.92 | 20.91 | 20.72 |
| Socioeconomic status: % of people in 4th quartile (most deprived) | 18.24 | 18.24 | 18.47 |
| % of people in rural areas | 23.93 | 23.91 | 23.86 |

\*Until September

### Variables

The main variable was the diagnosis of malignant neoplasms. Monthly incidence rates of malignant neoplasm per 10^5^ population were calculated.

Time-series analysis were performed globally, by sex, age groups (15-64 years old and >64 years old), type of neoplasm, socio-economic status and rurality. We assessed the socioeconomic status using the validated MEDEA deprivation index [29]. We categorised the MEDEA deprivation index into quartiles where 1st and 4th quartiles are least and most deprived areas, respectively. Rural areas were categorised separately and were defined as areas with less than 10,000 inhabitants and a population density lower than 150 inhabitants/km2.

We also performed the same time-series analysis for some related cancer diagnostic procedures with data available, such as mammograms, colonoscopies and chest radiographies.

### Statistical analysis

We calculated monthly malignant neoplasm and diagnostic procedures incidence rates per 10^5^ population as the number of new diagnoses or procedures divided by the number of alive populations assigned to ICS primary care practices and multiplied by 100,000.

We obtained the expected incidence for the study period using a time series regression, where the response variable was the incidence rate per 10^5^ inhabitants from 2014 to 2018, and the adjustment variables were the trend and seasonality of the time series. Dataset was divided into three sets: training set (from 2014 to 2018), validation set (2019) and analysis set (2020). We used the training set to adjust the model and validation set to test our method as a sensitivity analysis. We checked whether our method identified any excess or reduction of the monthly incidence rates in a regular year not affected by the COVID-19 pandemic. Finally, we projected the estimated time series to our analysis set.

The number of expected cancer diagnoses for each period was obtained multiplying the projected incidence by the population and dividing by 100,000. Excess or lack of malignant neoplasms and diagnostic procedures were defined as the number of observed minus expected diagnoses, estimated monthly and only for the periods where observed incidence were under the lower 95% confidence interval (95% CI) of the time-series. We also calculated the percentage of reduction as follows: (number of observed diagnoses - number of expected diagnoses) / number of expected diagnoses. This percentage was also calculated for the lockdown period and the post-lockdown period.

We calculated 95% CI for each estimate.

All analyses have been conducted using R, version 3.5.1 [30].

## Results

More than 4.9 million people older than 14 years were included in our analysis. The population structure in terms of age and gender has remained stable in the study period, as is shown in Table 1.

From January 2014 to September 2020, 273,379 new malignant neoplasms were registered in primary care. This represents a monthly average incidence per 10^5^ population of 72.4 during 2014-2018 period, 72.8 during 2019 and only 54.6 during 2020. From January to September 2020, 24,265 new cancers were recorded, of which 47.3% were women and 63.5% older than 64 years, with similar distribution to previous years. Regarding the type of cancer, non-melanoma skin cancers accounted for 24.8% of all new cancers diagnosed in 2020 (supplementary table 2 provides the number and monthly incidence average for each type of cancer and other variables for the three periods considered in our study).

**Table 2.** Estimated number (N) of underdiagnosed malignant neoplasms and diagnostic tests and percentage (%) of reduction compared with expected, overall and by lockdown and post lockdown periods.

|  | Total<br>(March - September) |  | Lockdown period<br>(March-June) |  | Post lockdown period<br>(July - September) |  |
| --- | --- | --- | --- | --- | --- | --- |
|  | N<br>(95% CI) | %<br>(95% CI) | N<br>(95% CI) | %<br>(95% CI) | N<br>(95% CI) | %<br>(95% CI) |
| **Total** | **8,766**<br>**(4,135 to 13,397)** | **34.0**<br>**(19.5 to 44.1)** | **6,262**<br>**(3,616 to 8,909)** | **40.5**<br>**(28.2 to 49.2)** | **2,504**<br>**(519 to 4,488)** | **24.3 (6.2 to 36.5)** |
| Between 15 and 64 | 2,238<br>(1,125 to 3,352) | 25.2<br>(15.4 to 32.1) | 1,997<br>(1,106 to 2,888) | 37.2<br>(24.7 to 46.2) | 241<br>(19 to 463) | 6.9<br>0.7 to 11.1) |
| Older than 64 | 5,829<br>(2,627 to 9,031) | 35.2<br>(19.6 to 45.7) | 4,094<br>(2,266 to 5,921) | 41.2<br>(27.9 to 50.3) | 1,736<br>(361 to 3,110) | 26.1<br>(6.9 to 38.8) |
| Women | 3,686<br>(1,618 to 5,754) | 31.6<br>(16.8 to 41.8) | 2,757<br>(1,576 to 3,939) | 39.2<br>(26.9 to 47.9) | 928<br>(42 to 1,814) | 20<br>(1.1 to 32.8) |
| Men | 5,089<br>(2,362 to 7,816) | 36.1<br>(20.8 to 46.4) | 3,509<br>(1,951 to 5,068) | 41.6<br>(28.3 to 50.7) | 1,580<br>(411 to 2,748) | 27.9<br>(9.1 to 40.2) |
| Skin (non-melanoma) | 3,118<br>(1,807 to 4,429) | 43.7<br>(31 to 52.4) | 2,346<br>(1,596 to 3,095) | 55.1<br>(45.5 to 61.8) | 773<br>(211 to 1,334) | 26.9<br>(9.1 to 38.8) |
| Melanoma | 243<br>(83 to 403) | 35.7<br>(18.2 to 44.6) | 202<br>(74 to 330) | 50.9<br>(27.5 to 62.9) | 41<br>(9 to 73) | 14.5<br>(4.8 to 19.3) |
| Colorectal | 568<br>(167 to 970) | 27.3<br>(11.0 to 36.7) | 478<br>(157 to 799) | 38.3<br>(16.9 to 50.9) | 91<br>(10 to 171) | 10.9<br>(1.7 to 15.9) |
| Prostate | 539<br>(189 to 890) | 33.3<br>(16.7 to 42.2) | 454<br>(174 to 735) | 45.2<br>(24.0 to 57.2) | 85<br>(15 to 155) | 13.8<br>(3.7 to 18.8) |
| Breast | 336<br>(173 to 500) | 17.1<br>(10.9 to 21.3) | 336<br>(173 to 500) | 27.8<br>(17.5 to 35.1) | 0 | 0 |
| Lung | 350<br>(49 to 650) | 20.0<br>(4 to 28.5) | 262<br>(37 to 487) | 25.4<br>(5.1 to 36.5) | 88<br>(13 to 163) | 12.2<br>(2.6 to 17.2) |
| Others | 2,585<br>(873 to 4,297) | 24.5<br>(10.7 to 33.2) | 1,829<br>(801 to 2,857) | 28.9<br>(16.2 to 37.1) | 755<br>(71 to 1,439) | 17.9<br>(2.2 to 27.4) |
| First quartile: urban least deprived | 2,092<br>(1,010 to 3,173) | 35.5<br>(21.8 to 44.3) | 1,636<br>(914 to 2,358) | 45.5<br>(31.8 to 54.6) | 456<br>(96 to 816) | 19.8<br>(5.5 to 28.7) |
| Second quartile | 963<br>(500 to 1,427) | 25<br>(16.4 to 30.6) | 818<br>(470 to 1,165) | 35.4<br>(25.4 to 42.0) | 146<br>(30 to 261) | 9.5<br>(2.5 to 13.8) |
| Third quartile | 1,557<br>(758 to 2,356) | 30.2<br>(18.8 to 37.5) | 1,354<br>(714 to 1,994) | 43.5<br>(28.9 to 53.2) | 203<br>(43 to 362) | 9.9<br>(2.7 to 14.3) |
| Fourth quartile: urban more deprived | 1,649<br>(760 to 2,539) | 35.7<br>(20.4 to 46.1) | 1,105<br>(596 to 1,613) | 39.9<br>(26.4 to 49.2) | 545<br>(164 to 925) | 29.4<br>(11.1 to 41.4) |
| Rural areas | 1,325<br>(686 to 1,965) | 21.3<br>(13.4 to 26.7) | 1,099<br>(620 to 1,579) | 29.9<br>(20.4 to 36.6) | 226<br>(66 to 386) | 8.8<br>(3.2 to 12.7) |
| Colonoscopy | 16,219<br>(10,836 to 21,603) | 49.8<br>(41.2 to 55.6) | 13,401<br>(9,812 to 16,990) | 66.1<br>(58.8 to 71.2) | 2,818<br>(1,024 to 4,613) | 22.9<br>(10.6 to 30.7) |
| Mammography | 15,099<br>(10,974 to 19,223) | 43.2<br>(39.5 to 45.6) | 15,099<br>(10,974 to 19,223) | 64.5<br>(56.9 to 69.8) | 0 | 0 |
| Chest radiography* | 12,638<br>(9,947 to 15,329) | 14.3<br>(9.3 to 22.1) | 12,638<br>(9,947 to 15,329) | 21.4<br>(14.2 to 31.7) | 0 | 0 |

\* In the case of chest radiography the results are the excess of radiographies instead of the underreported
\*\*In chest radiography the percentage represents the increase of radiographies instead of the reduction

Figure 1 shows the observed and estimated rates of monthly new malignant neoplasms diagnoses (with 95% CI) since January 2019. Observed incidences were as expected for 2019 and the first two months of 2020. Since March 2020, observed incidences of malignant neoplasms were significantly lesser than expected, for the whole population and by age groups.

**Figure 1.**
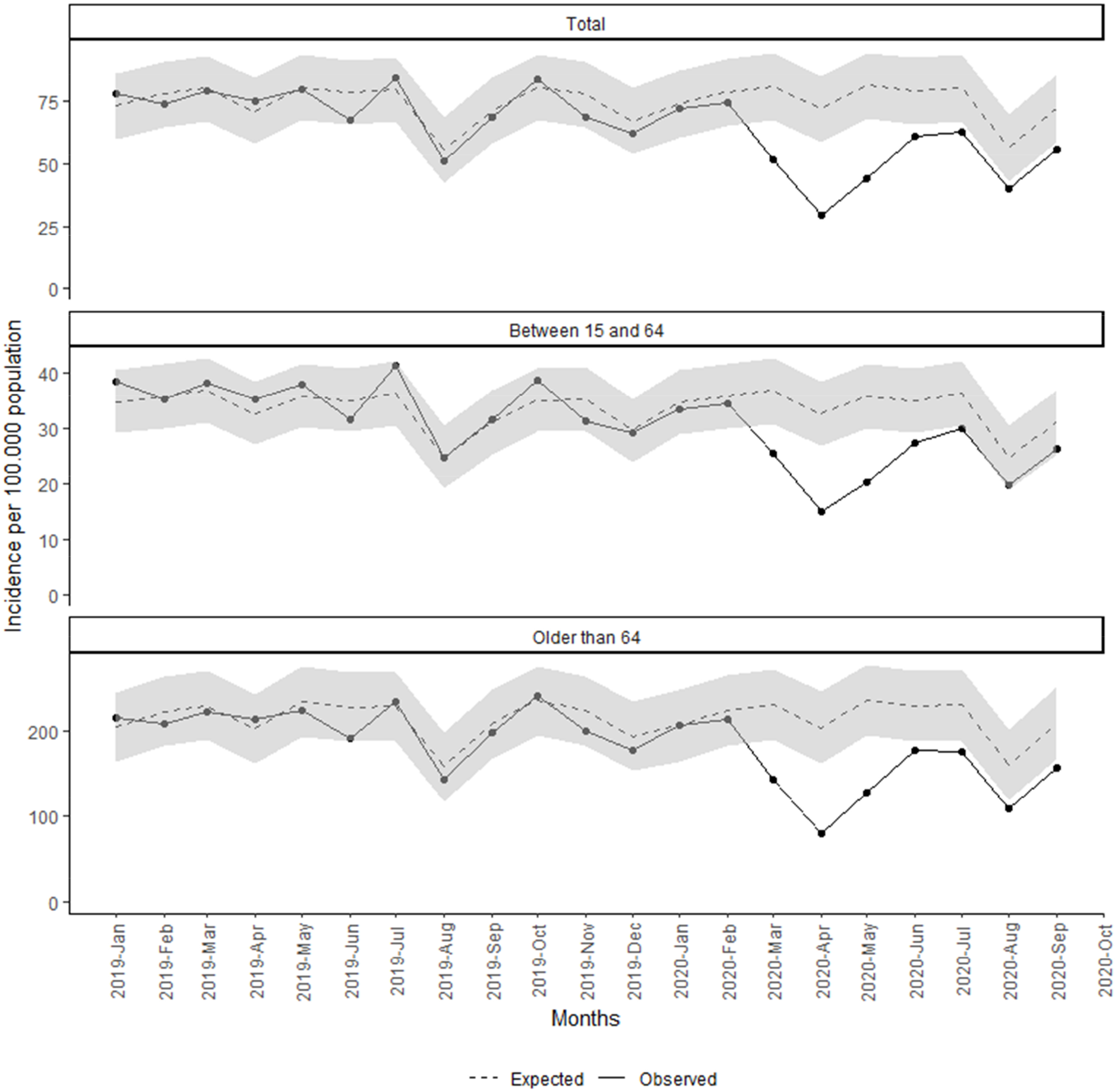
Monthly incidence of malignant neoplasm since January 2019. Total and by age group.

We estimated that the difference between expected and observed diagnoses accounted for 8,766 less cancers (95% CI: 4,135 to 13,397) from March to September. This represents a reduction of 34% of the diagnosis of malignant neoplasm compared to the expected. This reduction was greater during the lockdown period with 40.5% (95% CI: 28.2% to 49.2%) reduction compared with expected versus the 24.3% (95% CI: 6.2% to 36.5%) for the post-lockdown period (Table 2). The months with greater reduction were April, May and March with reductions of 59.3% (95% CI: 50% to 65.7%), 45.5% (95% CI: 34.8% to 53.2%) and 36% (95% CI: 23.2% to 45.1%), respectively (detailed expected cases, and number and percentages of cancer reductions by month are shown in supplementary table 3 stratified by age group, sex, socioeconomic status and type of cancer).

The estimated number of underdiagnosed malignant neoplasms and the percentage of underdiagnoses compared with the expected is presented stratified by age, sex, type of neoplasm, socio-economic status and rurality in Table 2. Population older than 64 years had more reduction than the population between 15 and 64 years, 35.2% (95% CI: 19.6% to 45.7%) versus 25.2% (95% CI: 15.4% to 32.1%). Regarding sex, reduction in men was greater than in women, 36.1% (95% CI: 20.8% to 46.4%) and 31.6% (95% CI: 16.8% to 41.8%) respectively (Figure 2 and Table 2).

**Figure 2.**
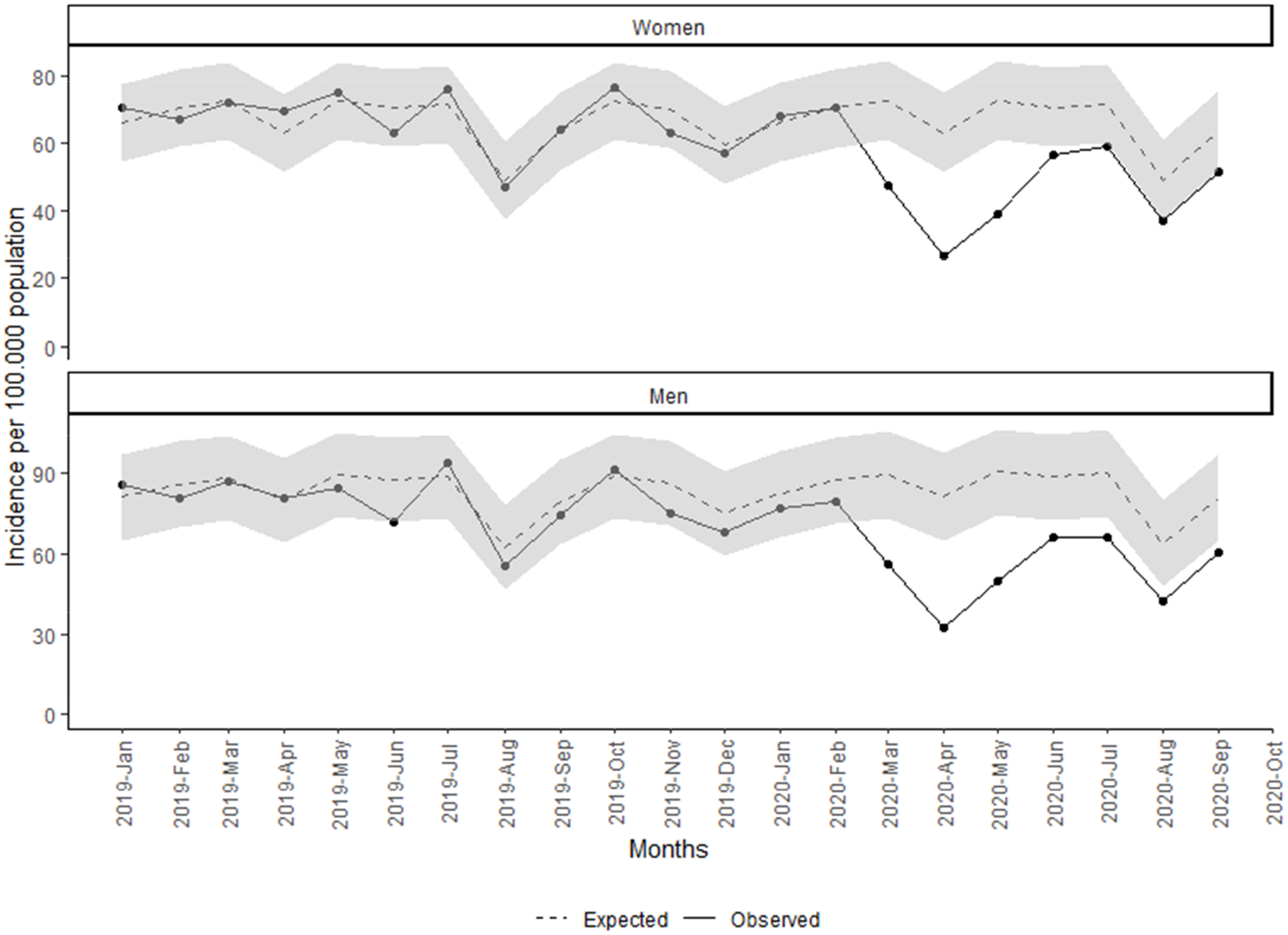
Monthly incidence of malignant neoplasm since January 2019 by sex.

Regarding the type of cancer, we found that in all cancers observed incidences were lesser than expected since March 2020 (Figure 3 and Table 2).This reduction was greater in April and since then incidence started to increase although still under the 95% CI until August, with big differences between the type of cancer. Non-melanoma skin cancers were those with higher reductions with 3,118 less cancers (95% CI: 1,807 to 4,429); followed by CRC with 568 less cancers (95% CI: 167 to 970), representing a reduction of 27.3% (95% CI: 11% to 36.7%); and prostatic cancers with 539 less cancers (95% CI: 189 to 890) and 33.3% reduction (95% CI: 16.7% to 42.2%) compared to the expected. Breast and lung new cancers were moderately reduced compared with other types of cancers, 17.1% (95% CI: 10.9% to 21.3%%) and 20% (95% CI: 4% to 28.5%) respectively. Incidences in lung cancer were under the 95% CI for fewer months than the other type of cancer, as shown in Figure 3. In addition, all the reduction of breast cancer was during the lockdown period.

**Figure 3.**
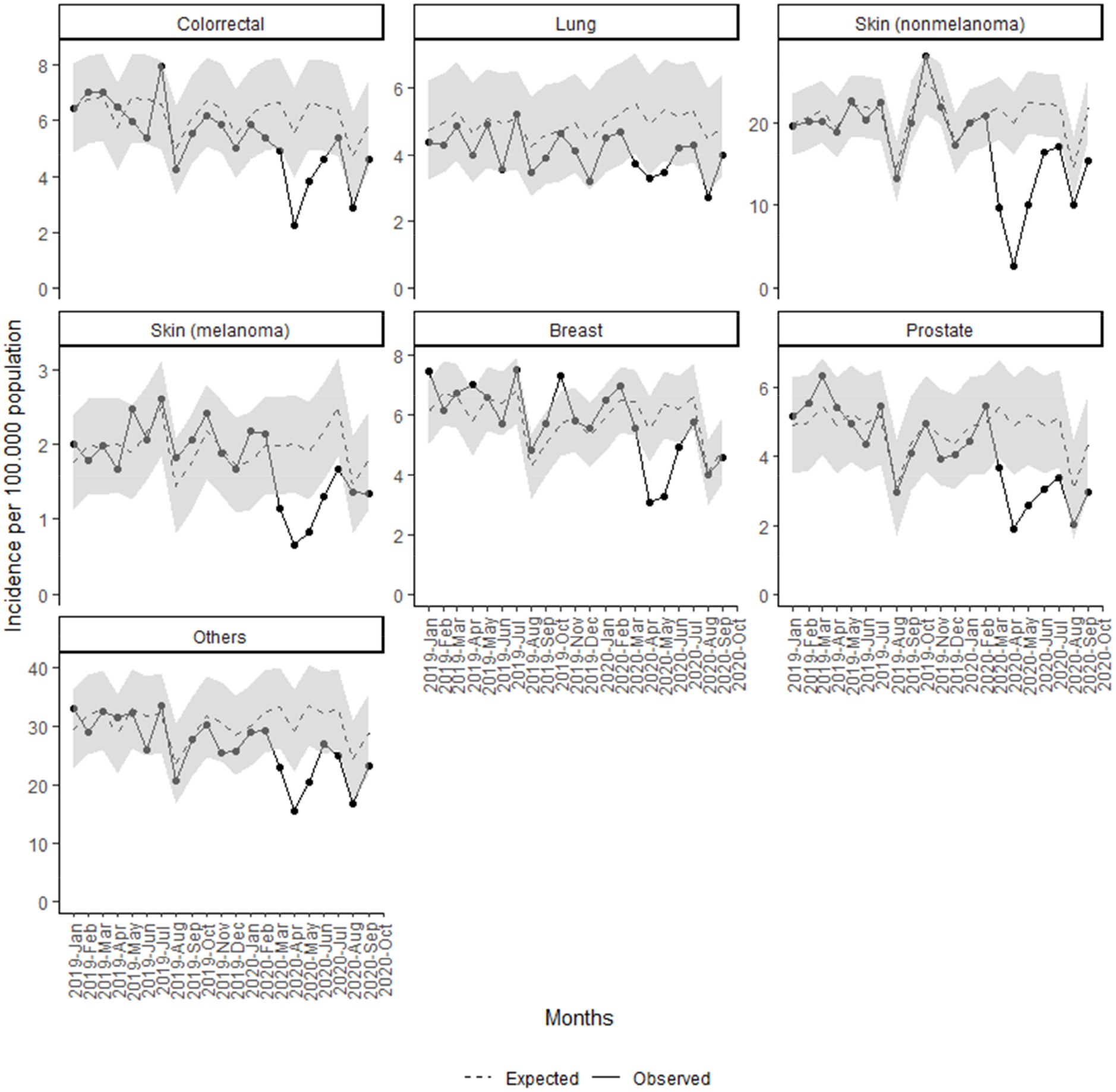
Monthly incidence of malignant neoplasm since January 2019 by type of cancer.

The same pattern and reduction of new malignant diagnosis were observed in all socio-economic status (Figure 4 and Table 2), although in rural areas it was the least with 21.3% (95% CI: 13.4% to 26.7%) reduction and only three months of statistically differences between observed and expected. Urban least and most deprived areas accounted for similar reduction although with some differences between periods. Least deprived areas had more reduction during the lockdown period (45.5% versus 39.9% in most deprived). Nonetheless, least deprived areas accounted for less reduction after the lockdown.

**Figure 4.**
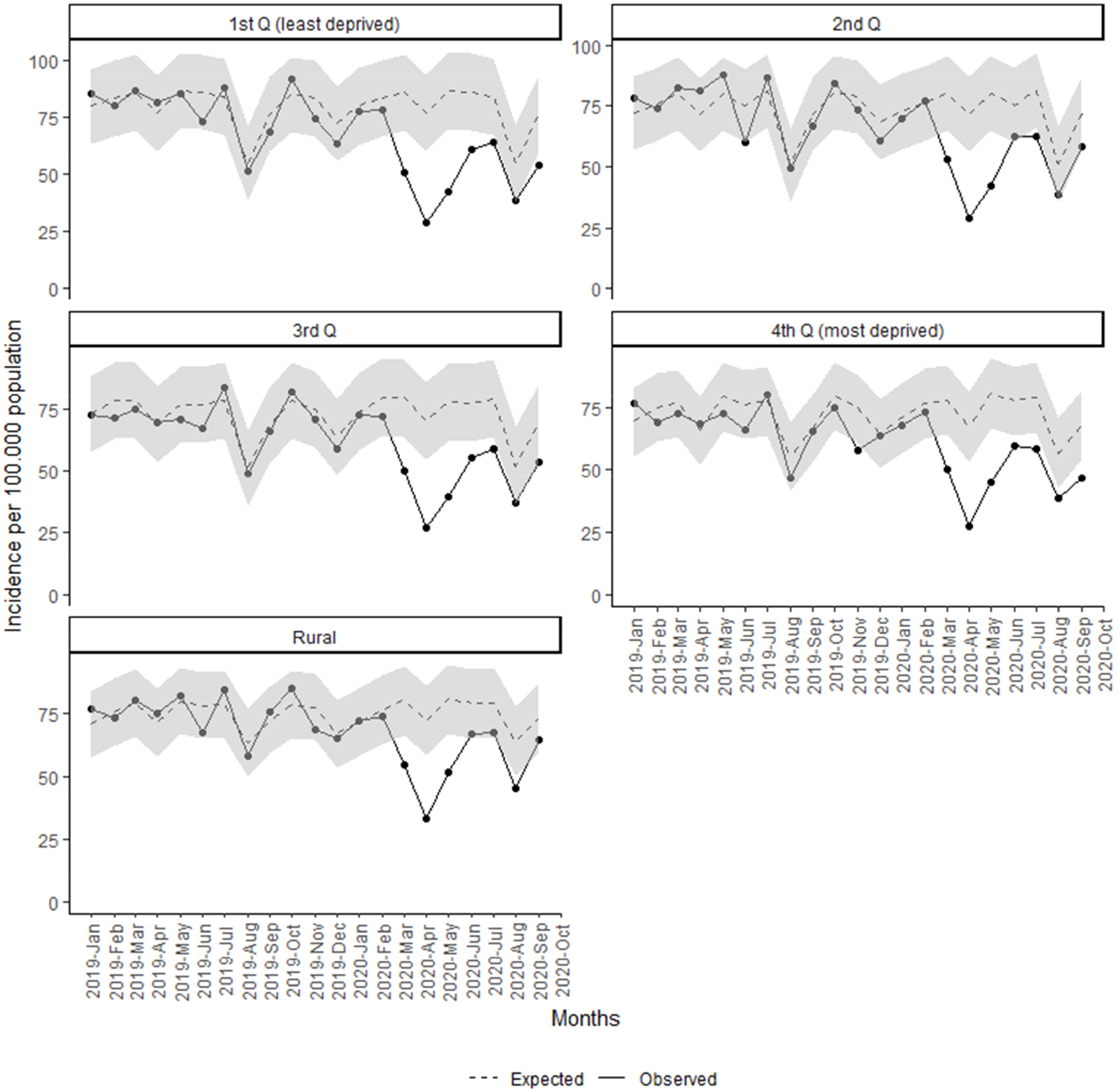
Monthly incidence of malignant neoplasm since January 2019 by socio-economic status and rurality.

Finally, Figure 5 shows the observed and estimated rates per 10^5^ population of monthly new mammograms, colonoscopies and chest radiographies. We observed a reduction of mammograms and colonoscopies during the same months as we observed for neoplasms and an excess of chest radiographies during April 2020. The reduction in procedures accounted for 16,219 less colonoscopies (95% CI: 10,836 to 21,603) and for 15,099 less mammograms (95% CI: 10,974 to 19,223) (Table 2). In addition, reduction of mammograms was only observed during the lockdown period.

**Figure 5.**
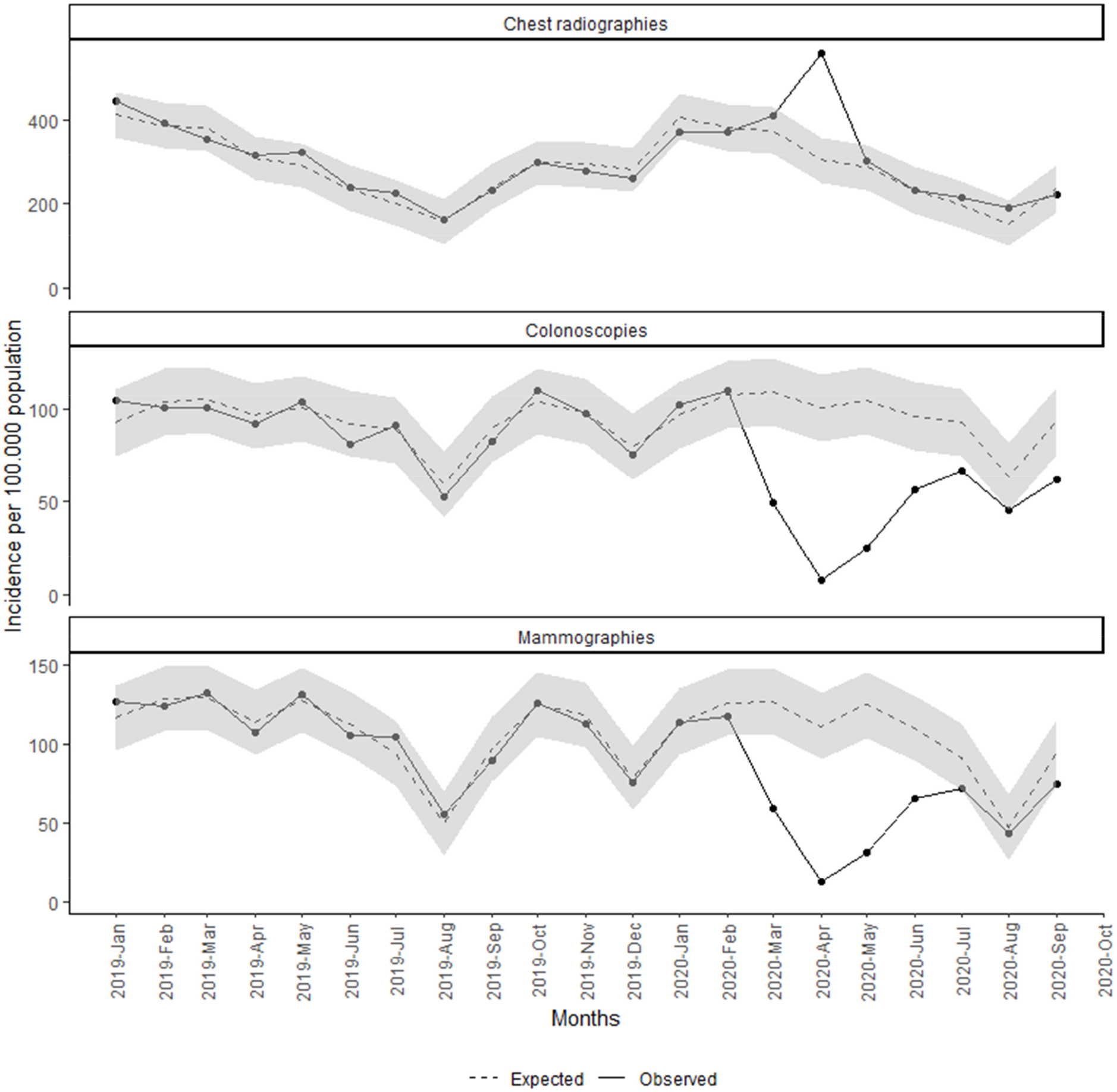
Monthly incidence of mammograms, colonoscopies and chest radiographies since January 2019.

## Discussion

Our study analyses the effects of the COVID-19 pandemic and related control measures on cancer diagnosis. We found a reduction of more than 8,700 new cancers since March 2020, that represents 34% less cancer than expected. This reduction was greater during the months of March, April and May, temporarily overlapping with the lockdown measures established in Spain in mid-March 2020 [7]. However, the reduction of new diagnoses of cancer extended beyond the end of the lockdown, especially for people older than 64 years, men, most deprived areas and some types of cancer; suggesting that there are other reasons that may caused this reduction. Other studies in different countries have observed reductions on cancer diagnosis and other diseases during the pandemic [12], with similar results. For example, in Italy, cancer diagnoses fell in 2020 by 39% compared with the average number recorded in previous years [31] and in Slovenia, it decreased 29% in April 2020 compared to the November 2019 - January 2020 average [23].

Our study reveals that almost all cancers have been affected. Nonetheless, the reduction of new cancer diagnoses ranged between 43.7% in non-melanoma skin cancers to 17.1% in breast cancers. In addition, breast cancer was only reduced during the lockdown period, also coinciding with the temporary stop of the population screening programmes. These differences between cancers are consistent with the impact on cancer diagnosis described in several studies, although these works are mainly focused on the first wave of COVID-19 period. In Italy, CRC has decreased by 62% and prostate cancer by 75%, having the greatest reductions; while breast cancer was moderately reduced (26%) [31]. In the UK, patients with new melanoma dropped by 67.1% in April 2020 compared with 2019, while lung cancer was the least altered (46.8% of reduction)[20]. In the Netherlands, researchers also found differences in cancer reductions between non-skin cancers (26%) and skin cancers (60%) [24].

The negative effect of the lockdown on the incidence of cancers appeared to occur across all ages, sex and socioeconomic status. Least deprived areas were those with greatest reductions during the lockdown period. This finding should be interpreted cautiously. There is evidence that least deprived areas have an increased access to healthcare [32]. During lockdown, with the stoppage of screening programmes and the halt of many diagnostic procedures in both public and private healthcare facilities, areas with better healthcare acces were more affected than those with already some problems of underdiagnosis due to preexisting inequalities [33, 34]. In addition, most deprived areas had a greater incidence of COVID-19 during those weeks and in consequence it is presumably that they have frequented more health services during lockdown. Nonetheless, it seems that after the lockdown period, most deprived areas had more reduction (29.4% vs 19.8%) and recovered worse than least deprived. This is concerning because at the end it is possible that reductions in most deprived areas last for more time than in least deprived, where people have access to private healthcare. These deprived areas should be areas of special attention and priority areas of governments’ actions and control measures to reduce inequalities that COVID-19 has exacerbated and highlighted.

This reduction was also observed in some cancer diagnostic procedures during the same period. Other studies found similar results in many countries [20]. In Argentina, colonoscopy has been reduced by 80% during the pandemic [35]. In Slovenia, a reduction of 48%, 75% and 42% were observed for X-rays, mammograms and ultrasounds [23]. In addition, as well as in our context, some of these reductions are related to a suspension or decline in cancer screenings services, with breast cancer screenings dropping by 89.2% and colorectal cancer screenings by 84.5% in some countries [20, 24, 36]. For example, a recent research in the Netherlands described that the temporary halt of national population screening programmes led to a fewer breast and CRC in age groups eligible for cancer screening programmes [37]. However, some studies pointed out that reduction of diagnostic procedures is also related to a decrease in cancer encounters, leading to a reduction in diagnostic suspicion [20].

Several reasons might explain our cancer diagnoses outcomes as well as the diagnostic procedures in the COVID-19 pandemic context. First, during the lockdown, all non-essential health activities were interrupted and authorities advised the population to stay at home. In addition, Spain was heavily hit by COVID-19 morbimortality in March to May, the health system collapsed and many diagnostic procedures were halted, except those used to diagnose some complications of COVID-19 such as chest radiography or thoracic CT scans. In our study, we found an excess of chest radiography probably due to an excess of COVID-19 related suspected pneumonia during April. But even after the lockdown restrictions, cancer incidence didn’t achieve the expected results, suggesting that measures to control the pandemic still have side effects in the diagnosis of cancer as well as other diseases. Second, some patients might have barriers to consulting a general practice, even after the lockdown. In Catalonia, the number of face-to-face visits has dropped drastically since March and has been replaced by telehealth [38], making it more difficult to appraise signs and symptoms and to initiate an investigation for a cancer diagnosis, especially for some populations, like more socioeconomic disadvantaged individuals who are less likely to engage in telemedicine [39]. Lastly, studies also suggested some reasons linked to health-seeking behaviour of the patients or the fear to be infected by SARS-CoV-2 [19, 24, 40]. This could partially explain the greatest reduction on non-melanoma skin cancer as some patients could consider some skin lesions as more demorables [41].

The 34% decrease in the incidence of new cancers in our study is considered to be concerning although the effects of these reductions are still uncertain, as the impact of the pandemic on cancer patients can only be reliably assessed after a reasonable time. However, some studies estimated an increase of the total years of life lost and other consequences[17, 22, 42, 43]. Because many cancers can advance rapidly, months without detection are likely to have highly detrimental effects on the stage of cancer at diagnosis. More advanced cancers stages are more difficult to treat successfully and, for many cancers, the stage of disease at diagnosis is related to survival and to cure rates [44], as well as the higher costs involved [36]. In addition, an increased demand for cancer procedures is expected to manage the backlog within routine diagnostic service [20], even though not observed yet in our study. These consequences differ according to tumour type [26]. For example, some tumors such as lung, pancreatic or breast cancers progress rapidly and require immediate diagnosis and treatment to prevent adverse outcomes [18, 45]. However, other slow-growing common cancers, such as prostate, cervical and non-melanoma skin cancers, are not so affected by the delay of diagnosis [46, 47]. Finally, another concern to consider is the psychological impact of cancer diagnosis delay in people who suffer from any symptoms that can be considered high risk of cancer [48].

Our study has some limitations. Firstly, we performed a time-series study with ecological data, but this approach does not allow to ensure a causal correlation between reduction of the incidence of malignant neoplasm and COVID-19 pandemic, although the reduction in the observed data around the start of the state of the alarm in March 2020 is clear. Secondly, as COVID-19 striked hard Spain with an excess of 44,000 deaths [49], some can argue that part of the reduction of the incidence of malignant neoplasms could be linked to the harvesting effect where some patients that could have been diagnosed had deceased. Even so, we observed a defect of the incidence of cancer in all age groups while the excess of deaths caused by COVID-19 was mostly in old people [49].

Despite the limitations, this study also has strengths. The data used were obtained from primary care EHR and are of good quality. Several studies have used the Catalan EHR to do useful research in real-world conditions [29, 50]. Moreover, our estimation used data from five years and we validated our method with 2019 diagnoses, where we didn’t find any excess or reduction of the incidence of cancer, strengthening our findings in 2020. In addition, our analysis extends the first wave of COVID-19-related effects and lasted for seven months in contrast to other studies focused on lockdown period only. We also assessed the socioeconomic differences, identifying the most affected areas that should be a priority during the following months. As ICS manages about 3 in every 4 practices in Catalonia, our results are generalizable and our method could be introduced in other settings that also use EHR. That way, our findings could be confirmed in other countries.

In conclusion, containing COVID-19 pandemic has been the health top priority during the last months. However, this situation caused a reduction in cancer incidence that suggests a delay in cancer diagnosis and, probably, future negative outcomes, still uncertain. We must continue to control the pandemic, but we also need to ensure that common causes of morbidity and mortality such as cancers are also taken into account when decisions are made. It can be assumed that during the coming months some partial or global lockdows will be put in place. Urgent policy interventions are necessary to mitigate the indirect effects of these measures on other diseases and some strategies must be designed in order to reduce this underdiagnosis of cancer. In addition, long-term studies should be performed in order to evaluate the future effects of this situation, such as a loss of survival or a shift toward a more advanced stage at diagnosis for some cancers.

## Supporting information

Supplementary tabble 3

## Data Availability

All data and analytical code are provided at: https://github.com/ErmengolComa/neos

## Patient and Public Involvement statement

This research was done without patient involvement. Patients were not invited to comment on the study design and were not consulted to develop patient relevant outcomes or interpret the results. Patients were not invited to contribute to the writing or editing of this document for readability or accuracy.

## Data sharing statement

All data and analytical code are provided at: https://github.com/ErmengolComa/neos

## Ethics approval

This study was done in accordance with existing statutory and ethical approvals from the Clinical Research Ethics Committee of the IDIAPJGol (project code: 20/172-PCV).

## Transparency declaration

The authors affirm that the manuscript is an honest, accurate, and transparent account of the study being reported; that no important aspects of the study have been omitted; and that any discrepancies from the study as planned have been explained

## Acknowledgments

We would like to acknowledge the efforts of all members of the SISAP team during the last months. We would also like to thank all the primary care healthcare professionals in Catalonia during these challenging times.

We also thank Ada Ferrer for her valuable contribution to language editing of the manuscript.

## Author contributions

All authors contributed to the design of the study, the interpretation of the results, and reviewed the manuscript. EC, FF and NM had access to the data, performed the statistical analysis and acted as guarantors. EC, CG, MM-C and NM wrote the first draft of the manuscript. All authors critically revised the manuscript. The corresponding author attests that all listed authors meet authorship criteria and that no others meeting the criteria have been omitted.

