## Supplementary tables 1 and 2 for "The impact of the COVID-19 pandemic and related control measures on cancer diagnosis in Catalonia: A time-series analysis of primary care electronic health records covering about 5 million people"

**Supplementary material**

**Supplementary Table 1.** ICD-10 codes and ICPC2 codes used to identify malignant neoplasms

| **Type of malignant neoplasm** | | **ICPC2** **Code** | **ICD-10 Code** |
| --- | --- | --- | --- |
| Colorectal | | D75 | C18% - C21% |
| Lung | | R84 | C33%, C34%, C46.5% |
| Skin non-melanoma | | S770 | C44%, C46.0, C46.1 |
| Melanoma | | S771 | C43% |
| Breast | | X76 | C50% |
| Prostate | | Y77 | C61% |
| Others | Stomach | D74 | C16% |
|  | Pancreas | D76 | C25% |
|  | Other digestive organs | D77 | C17%, C46.2, C46.4, C00% - C08%, C15%, C17%, C22%, C23%, C24%, C26%, C45.1, C46.2, C46.4, C48% |
|  | Brain and other parts of nervous system | N74 | C47%, C70%, C71%, C72% |
|  | Cervix uteri | X75 | C53% |
|  | Other female genital organs | X77 | C51%, C52%, C54%-C57% |
|  | Lymphoma | B72 | C81%-C86%, M31.2 |
|  | Leukemia | B73 | C90.1, C91%-C95% |
|  | Other hematopoietic and related tissue | B74 | C77%, C88%, C90, C90.0, C90.2, C90.3, C96%, C26.1, C37%, C46.3 |
|  | Bladder | U76 | C67% |
|  | Kidney and renal pelvis | U75 | C64%, C65% |
|  | Ureter and other urinary organs | U77 | C66%, C68% |
|  | Thyroid | T71 | C73% |
|  | Other respiratory organs | R85 | C09%-C14%, C30%-C32%, C39%, C45.0 |
|  | Bone and articular cartilage | L71 | C40%, C41%, C46.1, C49% |
|  | Breast and genital organs in men | Y78 | C50, C50.1, C50.9, C60%, C62%, C63% |
|  | Ear | H75 | C30.1 |
|  | Eye | F74 | C69% |
|  | Heart | K72 | C38.0, C45.2 |
|  | Pregnancy | W72 | C58 |
|  | Miscellanea | A79 | C14%, C30%, C38%, C39%, C45, C45.7, C45.9, C46, C46.7, C46.9, C4A%, C76%, C77%, C78%, C79%, C7A%, C7B%, C80 |

**Supplementary Table 2. Number and monthly incidence average of new cancers for each study period stratified by type of cancer, sex, age groups, and socieoconomic status**

| Variable | Period | Number of cancer diagnoses | Monthly incidence average |
| --- | --- | --- | --- |
| Total | 2014-2018 | 206,470 | 72.4 |
|  | 2019 | 42,644 | 72.8 |
|  | 2020 | 24,265 | 54.6 |
| Between 15 and 64 | 2014-2018 | 75,376 | 33.8 |
|  | 2019 | 15,639 | 34.4 |
|  | 2020 | 8,863 | 25.7 |
| Older than 64 | 2014-2018 | 131,094 | 210.1 |
|  | 2019 | 27,005 | 205.9 |
|  | 2020 | 15,402 | 154.9 |
| Women | 2014-2018 | 96,339 | 66.1 |
|  | 2019 | 19,992 | 66.7 |
|  | 2020 | 11,484 | 50.6 |
| Men | 2014-2018 | 110,131 | 78.9 |
|  | 2019 | 22,652 | 79.1 |
|  | 2020 | 12,781 | 58.9 |
| 1st Q (least deprived) | 2014-2018 | 48,672 | 79.6 |
|  | 2019 | 9,858 | 77.5 |
|  | 2020 | 5,299 | 55.0 |
| 2nd Q | 2014-2018 | 31,326 | 72.7 |
|  | 2019 | 6,581 | 73.7 |
|  | 2020 | 3,698 | 54.6 |
| 3rd Q | 2014-2018 | 41,471 | 69.2 |
|  | 2019 | 8,475 | 69.7 |
|  | 2020 | 4,766 | 51.8 |
| 4th Q (most deprived) | 2014-2018 | 35,656 | 67.7 |
|  | 2019 | 7,331 | 67.9 |
|  | 2020 | 4,261 | 52.0 |
| Rural | 2014-2018 | 49,345 | 72.0 |
|  | 2019 | 10,399 | 74.3 |
|  | 2020 | 6,241 | 58.8 |
| Colorrectal | 2014-2018 | 19,935 | 7.0 |
|  | 2019 | 3,575 | 6.1 |
|  | 2020 | 1,963 | 4.4 |
| Lung | 2014-2018 | 11,630 | 4.1 |
|  | 2019 | 2,463 | 4.2 |
|  | 2020 | 1,716 | 3.9 |
| Skin (nonmelanoma) | 2014-2018 | 55,398 | 19.4 |
|  | 2019 | 11,948 | 20.4 |
|  | 2020 | 6,024 | 13.6 |
| Skin (melanoma) | 2014-2018 | 5,414 | 1.9 |
|  | 2019 | 1,191 | 2.0 |
|  | 2020 | 622 | 1.4 |
| Breast | 2014-2018 | 18,600 | 6.5 |
|  | 2019 | 3,721 | 6.3 |
|  | 2020 | 2,195 | 4.9 |
| Prostate | 2014-2018 | 13,869 | 4.9 |
|  | 2019 | 2,790 | 4.8 |
|  | 2020 | 1,447 | 3.3 |
| Others | 2014-2018 | 81,624 | 28.6 |
|  | 2019 | 16,956 | 28.9 |
|  | 2020 | 10,298 | 23.2 |
